# The MidPIC study: Midwives’ knowledge, perspectives and learning needs regarding preconception and interconception care

**DOI:** 10.1101/2023.07.31.23293432

**Authors:** Zoe Bradfield, Emily Leefhelm, Sze-Ee Soh, Kirsten I Black, Jaqueline A Boyle, Lesley Kuliukas, Cheryce Harrison, Caroline SE Homer, Rachel M Smith, Helen Skouteris

## Abstract

**Background:** Preconception and interconception care improves the health outcomes of women and communities.

**Problem:** Little is known about how prepared and willing Australian midwives are to provide preconception and interconception care.

**Aim:** The aim of this study was to explore midwives’ knowledge, perspectives and learning needs, and barriers and enablers to delivering preconception and interconception care.

**Methods:** We conducted a cross-sectional exploratory study of midwives working in any Australian maternity setting. An online survey was administered that included items measuring midwives’ self-rated knowledge; education needs and preferences; attitudes and perceptions towards the pre and interconception care; and views on future service and workforce planning.

Quantitative data were analysed descriptively, and demographic characteristics (e.g., years of experience, model of care) associated with knowledge and attitudes regarding pre- and inter-conception care were examined using univariate logistic regression analysis. Qualitative data were captured through open-ended questions and analysed using inductive content analysis.

**Findings:** We collected responses from (*n*=338) out of 355 midwives who were eligible for this study working across all Australian models of care (completion rate 96%). Most participants (*n*=290; 85%) rated their overall knowledge about pre and interconception health for women as excellent, above average or average. The only variable associated with overall knowledge was years of experience, with participants more than 11 years of experience more likely to report above average to excellent knowledge (OR 3.11; 95% CI 1.09, 8.85). The majority (*n*=257; 76%) were interested in providing pre and interconception care more regularly within their role. Low prioritisation in service planning/budgeting was the most frequently selected barrier to providing preconception and interconception care.

**Implications:** Findings revealed that midwives are prepared and willing to provide preconception and interconception care. Pre and post registration professional development; service and funding reform; and policy development are critical to enable Australian midwives’ provision of pre and interconception care.

## Introduction

Preconception care (PCC) and interconception care (ICC) refers to biomedical, social, and behavioural interventions and health counselling that occurs before or between pregnancies (1). PCC and ICC (PICC) improves health outcomes of women, newborns, children and their communities by promoting, maintaining and enhancing the health of women before a first, or subsequent pregnancy (2). All women have a right to healthcare that optimises their health, including preconception care as this enhances social capital (3) and strengthens agency against gender-based violence and economic inequality (4).

PICC is a critical mechanism to optimise the health of women during the reproductive life stage. There is a plethora of evidence supporting the association between healthy preconception states and reduced risk of adverse pregnancy outcomes, for example, physical activity before pregnancy is associated with a lower risk of preeclampsia (5), and ‘Mediterranean-style’ diets in the three years before pregnancy has been shown to reduce the risk of gestational diabetes (6). The preconception and interconception periods are also a critical time to address reproductive health conditions with long-term non-reproductive complications, for example, polycystic ovary syndrome in women of reproductive age has been shown to present a 1.3-fold risk of cardiovascular disease in later life (7), Additionally, women with a history of gestational diabetes have a significantly increased risk of developing type 2 diabetes post-pregnancy (8), First Nations Australian women and women who experience social disadvantage are 1.3 and 1.6 times respectively, more likely to develop gestational diabetes than other groups, which presents a higher risk of developing type 2 diabetes within 2.5 years of giving birth (9, 10).

Increasing evidence also supports the importance of preconception health on the health of future infants and children (11). Globally, the First 1000 days (12, 13) and First 2000 days (14–16) Frameworks reaffirm the lifelong impact that preconception and interconception health states have on infant and childhood development emphasising the need to prioritise accessible PICC for all individuals of reproductive age. The 2021 State of the World’s Midwifery Report revealed that midwives are capable of providing 90% of the world’s sexual and reproductive health (SRH) needs (17). While SRH across the reproductive life course is within the scope of midwifery practice (18, 19), midwives in Australia largely practice in settings related to pregnancy, birth and the six week postnatal period. Supporting midwives to fulfil their scope in the provision of PICC outside of the pregnancy to 12 weeks postpartum period presents an opportunity to improve the health states of women, babies, and communities (17).

Recently, Australian research has focused on the potential of primary health care nurses to provide PCC in primary health settings (20), with lack of time and knowledge cited as the key barriers to fulfilling this role (21). Research on midwives’ role in PICC has been limited. An older study conducted in 2006 in the Netherlands explored Dutch midwives’ perspectives on PCC and found that the traditional organisation of antenatal care restricted midwives’ access to women before pregnancy (22). Recent research conducted in Australia with midwives working in a tertiary setting revealed that whilst the midwives were keen and able to provide SRH care, they identified a strong desire for further education including in PICC (23). While midwives have been included in Australian efforts to improve the knowledge of nurses and midwives to promote preconception health (24), the voice of midwives in PICC evidence, service provision and policy is limited, submissions noting this gap have formed part of recent midwifery advocacy work in Australia (25).

The aim of this research was to explore midwives’ knowledge, perspectives, learning needs and their perceived barriers and enablers to delivering PICC. Generating this new knowledge is a pivotal step towards enabling scope-fulfilment for midwives to provide equitable, woman-centred preconception care to women across Australia, with learnings potentially generalisable internationally.

## Methods

Cross-sectional designs have recognised utility for collecting and measuring data at discrete points in time and provide important access to benchmarking data for new fields of discovery (26). Given the aforementioned paucity of evidence, a cross-sectional design was considered an ideal approach to address the study aim of exploring midwives’ knowledge, perspectives and learning needs regarding PICC, and in identifying enablers and barriers to the provision of PICC. Human Research Ethical approval was granted through Curtin University (HRE2022-0565). All participants provided informed consent prior to completing the survey.

## Study setting and context

There are 26,350 midwives employed in Australia (27). Of these, 23,642 operate in clinical roles and 2,708 are employed in non-clinical settings such as teaching, administration and research; 72.7% work in metropolitan areas (27). Most midwives are employed by public hospitals and provide care in a variety of models such as standard care (randomly allocated midwife provides care to a different woman at each visit); and continuity care (midwife partners with a woman and is her lead maternity carer throughout the childbearing experience) As an autonomous, independent profession, graduates of midwifery courses come from a range of life and professional backgrounds and are able to work to full scope on registration. An additional endorsement to prescribe medications and order diagnostic tests is available, post initial registration, for midwives who undertake further formal studies. These midwives are known as ‘endorsed midwives’ and often work in private practice or in primary care settings such as community clinics. Midwives also work in private obstetric-led services; in these models, midwives have irregular antenatal and postnatal contact with the women who have contracted with a private obstetrician. There are 1,028 endorsed midwives in Australia, this figure has doubled in the last two years and is expected to continue this trajectory of growth in the coming two years (28). It is common for individuals to practice a variety of professions prior to becoming a midwife, some will continue professional practice in a range of areas in tandem with midwifery practice. The rich tapestry and breadth of experience within the Australian midwifery workforce is relevant to the setting and context of this study.

### Survey design

Owing to the novel nature of this research, there were no existing validated tools to collect information regarding midwives’ knowledge, perspectives and learning needs in relation to pre and interconception care. The survey tool was developed by the research team who have content and research expertise. Discipline backgrounds of the researchers include including midwifery, obstetrics, women’s health, nursing, psychology, and public health. Several authors (ZB, KB, JB, HS) are also members of the recently formed International Core Indicators for Preconception Health and Equity (iCIPHE) Alliance comprising of representatives from more than 45 institutions and 20 countries globally (29).

The survey was designed in five parts, including: i) demographic data; ii) Likert questions regarding midwives’ self-rated knowledge; iii) midwives’ education needs and preferences; iv) Likert responses regarding midwives’ attitudes and perceptions towards the provision of PICC; and iv) questions regarding barriers and enablers of PICC service delivery (Supplementary File 1). Finally, there was opportunity to leave free text comments for a range of questions throughout the survey enabling the provision of descriptive responses. Despite international publications and guidelines (30) regarding PCC at the time of survey development, there was a lack of globally agreed core indicators and high-quality guidelines regarding PICC to support survey development. As such, national priorities outlined in a recent Delphi Study were used to support five key domains of PICC namely: optimising health behaviours; addressing pre-existing health conditions; achieving a healthy weight; optimising reproductive health; and optimising mental health (31). These domains align with contemporary international consensus (32, 33).

### Recruitment, sampling and data collection

Recruitment for this online survey was via Australian professional midwifery association newsletters, social media sites, QR codes at national midwifery conferences and through professional association member emails from 12^th^ October 2022 to 30^th^ December 2022. As an exploratory study, sampling was not driven by statistical calculation nor theoretical saturation; rather by the purposive convenience sampling strategy adopted.

Data were collected via an anonymous online survey hosted on Qualtrics (November 2022) a secure, encrypted, online survey platform. All surveys were completed via an anonymous generic link available on recruitment flyers.

### Data analysis

Survey data were analysed descriptively using Stata/IC 16.0 (StataCorp College Station, Texas, USA). Demographic characteristics (e.g., professional qualification, years of experience, model of care) associated with knowledge and attitudes regarding pre- and inter-conception care were examined using univariate logistic regression analysis. Model findings were reported as odds ratio (OR) with 95% confidence intervals (CIs), and *p*≤0.05 was considered statistically significant. Listwise deletion method was used to adjust for missing data.

Open-ended survey responses were coded generating categories using an inductive content analysis (ICA) approach; this is a useful approach when there are limited data available of the phenomenon under study as was the case here (34). The methodology supports the presentation of meaningful descriptions and abstractions of the raw data situated in individuals’ context; and follows three main steps of data reduction, grouping and the formation of concepts. Initially, open codes formed subcategories which were further grouped into categories and main categories (35). Verbatim quotes are presented to support the presentation of categories and main categories and are italicised in text with the participant number to evidence the range of responses.

## Quantitative results

### Survey responses

There was a total of 355 valid responses where participants proceeded to the survey questions, with 11 participants (3%) exiting the survey prematurely without providing any responses to the demographic section of the survey. A further six participants (2%) did not provide any responses to the content questions in the survey (i.e., questions relating to knowledge, education, attitudes and service planning). There were no significant differences in qualification type (Χ^2^= 1.03; *p*=0.960), years of experience (Χ^2^= 3.26; *p*=0.353) and region of practice (Χ^2^= 2.56; *p*=0.464) between those who responded to the content questions and those who did not. As such, only participants who provided data to both the demographic (part i) and content sections (parts ii-v) of the survey were included in this set of analysis (n=338).

### Participant characteristics

Around half (55%) of midwife participants held a registration as a registered nurse and 19% indicated registration as endorsed midwives (Table 1). The majority (96%) were female, born in Australia (72%) and completed their midwifery education in Australia (83%). Around one quarter (24%) held a graduate diploma as their highest degree and 23% had completed either a masters or PhD program. There were relatively even proportions of participants with <5 years (27%), 6-10 years (23%), 11-20 years (20%) and over 21 years’ (28%) of experience as a midwife. Over half (57%) worked in metropolitan areas, 28% in regional areas and 14% in rural or remote areas. Just under half (44%) worked in standard public non-continuity models, 15% in public hospital-based continuity models and 8% indicated they worked as a privately practicing midwife.

**Table 1.** Participant demographics.

|  | **All participants**<br>**n = 338** |
| --- | --- |
| Professional qualification* |  |
| Midwife | 338 (100) |
| Registered nurse | 186 (55) |
| Endorsed midwife | 65 (19) |
| IBCLC Lactation Consultant | 24 (7) |
| Maternal Child Health/Child Health qualification | 20 (6) |
| Other professional qualifications | 31 (9) |
| Gender identity |  |
| Female | 325 (96) |
| Male | 5 (1) |
| Non-binary | 3 (<1) |
| Prefer not to say | 2 (<1) |
| Australian-born |  |
| Yes | 245 (72) |
| No | 72 (21) |
| Language other than English |  |
| Yes | 23 (7) |
| No | 305 (90) |
| Completed midwifery education in Australia |  |
| Yes | 280 (83) |
| No | 42 (12) |
| Highest qualification level |  |
| Diploma/hospital-based training | 22 (7) |
| Bachelor's degree | 122 (36) |
| Graduate certificate | 35 (10) |
| Graduate diploma | 81 (24) |
| Masters | 65 (19) |
| PhD/Doctoral | 12 (4) |
| Year of experience as midwife |  |
| <5 years | 91 (27) |
| 6-10 years | 79 (23) |
| 11-20 years | 69 (20) |
| >21 years | 95 (28) |
| Australian jurisdiction for work |  |
| NSW | 62 (18) |
| VIC | 66 (20) |
| QLD | 56 (17) |
| SA | 16 (5) |
| WA | 110 (33) |
| TAS | 4 (1) |
| NT | 6 (2) |
| ACT | 15 (4) |
| Region of work |  |
| Metropolitan | 193 (57) |
| Regional | 93 (28) |
| Rural | 36 (11) |
| Remote | 11 (3) |
| Model of care |  |
| Standard public system, non-continuity model | 149 (44) |
| Standard private system, non-continuity model | 19 (6) |
| Public, hospital-based continuity model | 50 (15) |
| Public, community-based continuity model | 18 (5) |
| Private clinic/system, community-based continuity model | 13 (4) |
| Privately practicing midwife | 27 (8) |
| Education | 14 (4) |
| Research/policy/management | 17 (5) |
| Other (e.g., sexual health service, medical retrieval and Aboriginal Health Service) | 28 (8) |

213 All data reported as n(%) unless stated otherwise.
Not all cells sum to 100% owing to missing data.
\*Percentages add up to more than 100% where participants could select more than one option.

### Self-rated knowledge about pre and interconception health

Most participants (85%) rated their overall knowledge about pre and interconception health for women as excellent, above average or average (Fig 1). This differed to overall knowledge about pre and interconception health for men/partners, where a higher proportion of participants (40%) reported their overall knowledge as below average, poor or none.

**Fig 1.**
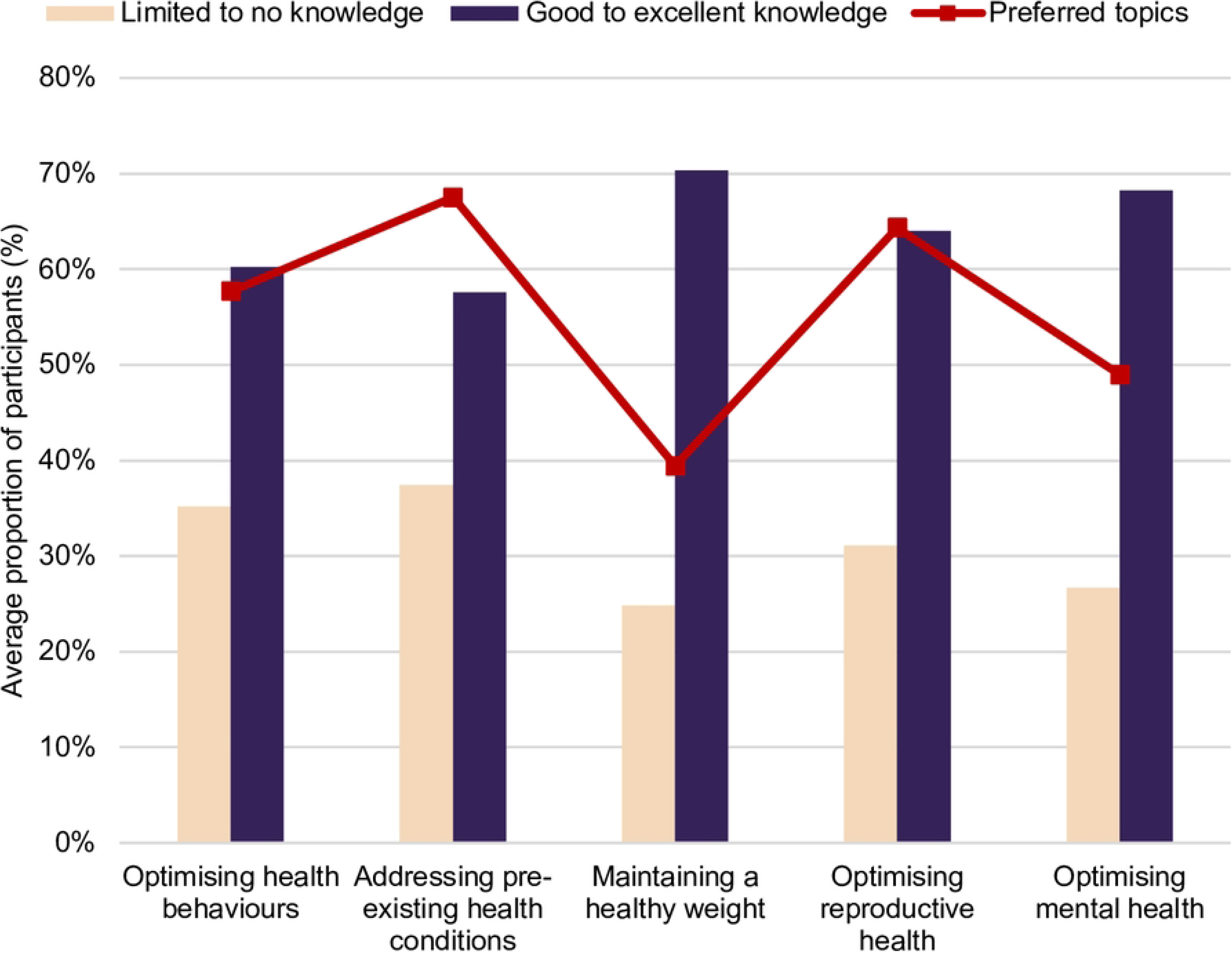
Midwives’ self-reported knowledge of pre and interconception care topics and preferred topics for further education. While overall reported topic knowledge was high, responses within each subcategory did reveal variable knowledge levels (Fig 2). Around half (52%) of participants reported no or limited working knowledge on screening for haemoglobin disorders, and despite 70% indicating good to excellent knowledge on healthy weight maintenance, 44% reported no or minimal working knowledge on disordered eating/eating disorders.

Knowledge on pre-existing health conditions had the lowest average proportion that reported good to expert working knowledge in this area, and unsurprisingly, managing pre-existing health conditions was selected by the highest proportion of participants (69%) as a desired topic for further education. A similar proportion (66%) also indicated a preference for education on optimising reproductive health (Fig 1). This was unsurprising given that less than half (44%) of the sample reported good to expert knowledge on abortion screening, discussion and assessment (Fig 2).

**Figure 2.**
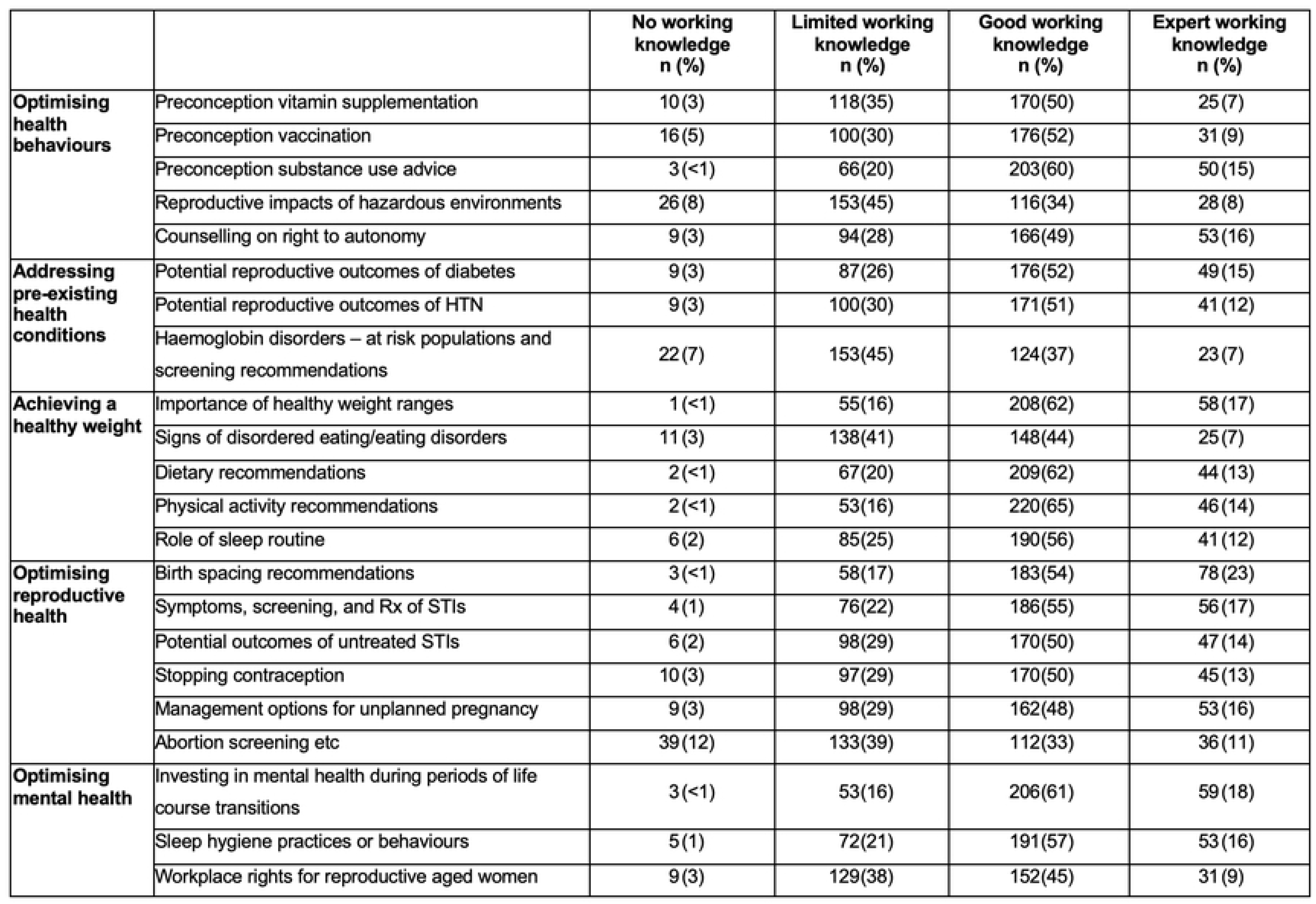
**Participants’ self-reported knowledge of PICC topic area subcategories (*n*=338).**

### Education needs and preferences

The top three preferred education formats were online e-learning courses (72%), face-to-face training within workplaces (46%) and online webinars (45%) (Fig 3).

**Fig 3.**
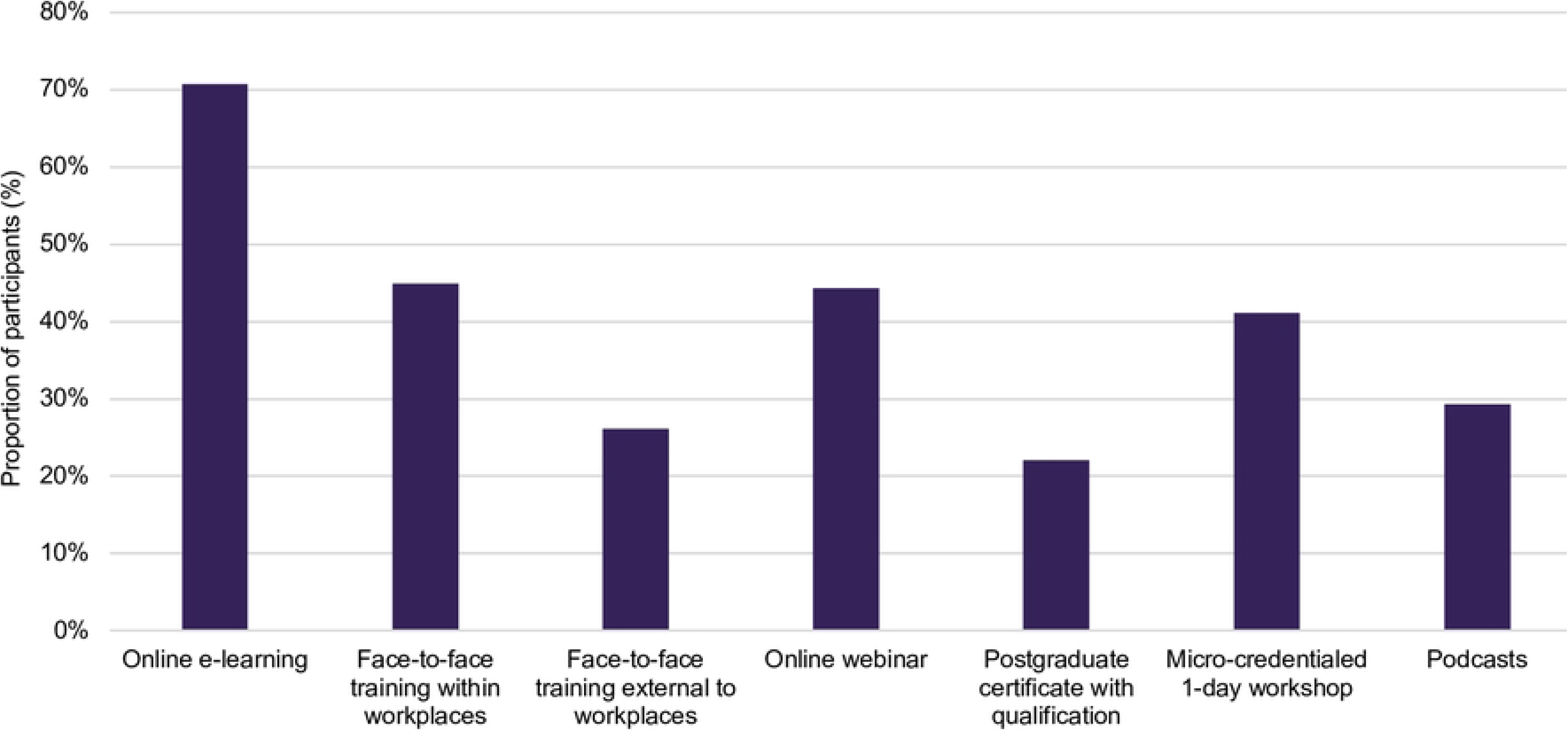
**Midwives’ preferred formats for PICC education.** *Percentages add up to more than 100% as participants could select more than one option

### Attitudes and perceptions towards the provision of PICC

Overall, participants indicated similar attitudes towards the included statements on PICC (Fig 4). The majority agreed or strongly agreed that pre and interconception care should be provided for all people of reproductive age (87%), that PICC is within the midwifery scope of practice (87%) and that they often encounter health states that could be managed before pregnancy (88%).

**Fig 4.**
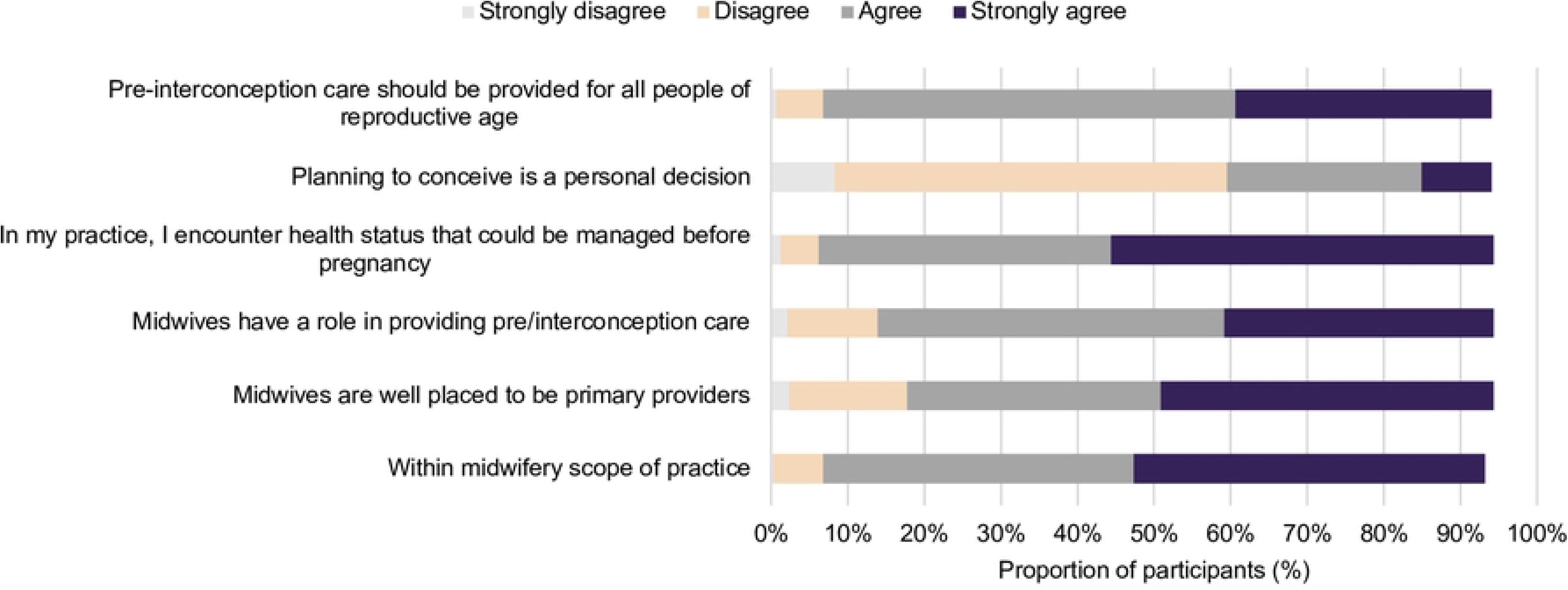
**Midwives’ attitudes surrounding PICC.**

### Barriers and enablers of PICC service delivery

A lack of prioritisation in service planning/budgeting was most frequently selected as barrier to providing PICC, indicated by 62% of participants (Fig 5). This was followed by time and staffing constraints (40%) and lack of prioritisation by healthcare professionals (40%).

**Fig 5.**
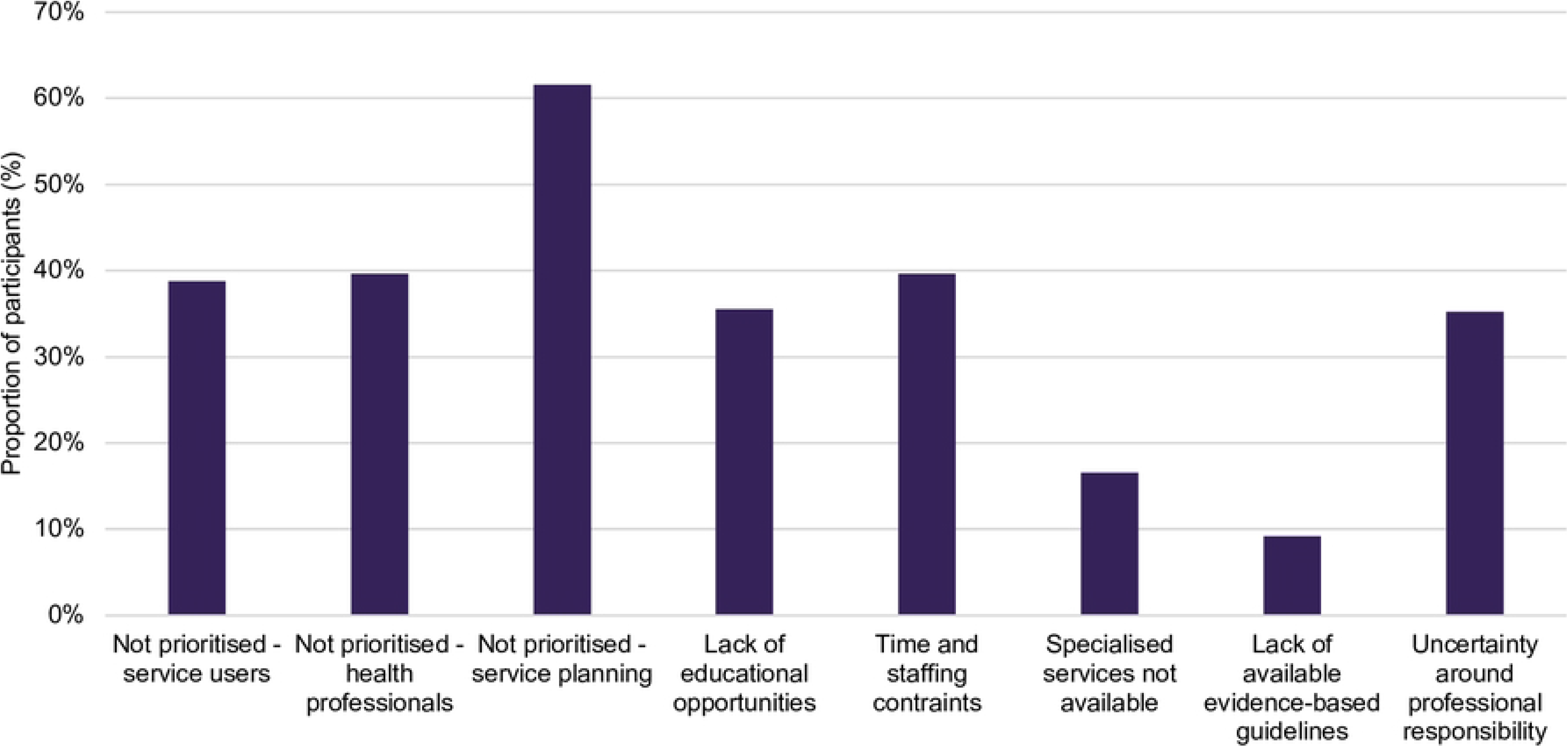
**Midwives’ self-reported barriers to establishing midwives’ role in the provision of PICC.** *Percentages add up to more than 100% as participants could select more than one option

Other barriers to establishing midwives’ role in PICC included lack of midwife presence in settings which would enable greater opportunity and provision of pre- or inter-conception care. Addressing this barrier would help address another identified barrier of midwives often only seeing women once they were pregnant.

When asked about service planning to enable PICC provision, 76% of midwives indicated that they ‘would’ and 15% reported they ‘may’ be interested in providing PICC more regularly as part of their current roles. When asked about the setting of work, 63% indicated they ‘would’ and 12% said they ‘may’ be interested in working in a community setting in order to provide PICC.

### Factors associated with overall knowledge about pre or interconception health

The only variable associated with overall knowledge about pre or interconception health for women was years of experience (Table 2). Participants with more than 11 years of experience were more likely to report above average to excellent knowledge (OR 3.11; 95% CI 1.09, 8.85). For knowledge about pre and interconception health for men/partners, this association was limited to midwives with more than 21 years of experience (OR 2.20; 95% CI 1.18, 4.11).

**Table 2.** Characteristics of participants associated with overall knowledge of PICC.

|  | Odds ratio [OR], 95% CI |  |
| --- | --- | --- |
|  | Overall knowledge about PICC for women | Overall knowledge about PICC for men/partners |
| Professional qualification |  |  |
| Midwife | 1 | 1 |
| Registered nurse | 1.41 (0.74, 2.64) | 1.39 (0.89, 2.17) |
| Professional qualification |  |  |
| Endorsed midwife | 1 | 1 |
| Endorsed midwife and registered nurse | 2.14 (0.36, 12.63) | 2.49 (0.87, 7.12) |
| Years of experience |  |  |
| <5 years | 1 | 1 |
| 6-10 years | 1.13 (0.52, 2.45) | 1.33 (0.62, 2.08) |
| 11-20 years | **3.11 (1.09, 8.85)** | 1.57 (0.83, 3.00) |
| >21 years | **3.10 (7.23, 7.83)** | **2.20 (1.18, 4.11)** |
| Highest qualification |  |  |
| Hospital-based diploma | 1 | 1 |
| Bachelor’s degree | 0.70 (0.19, 2.59) | 1.00 (0.36, 2.78) |
| Graduate certificate | 1.68 (0.31, 9.20) | 1.86 (0.56, 6.13) |
| Graduate diploma | 1.26 (0.31, 5.13) | 1.25 (0.44, 3.59) |
| Masters or PhD | 1.56 (0.37, 6.60) | 1.40 (0.48, 4.05) |
| Region of practice |  |  |
| Metropolitan | 1 | 1 |
| Non-metropolitan | 1.31 (0.68, 2.53) | 1.19 (0.75, 1.87) |
| Model of care |  |  |
| Standard public system | 1 | 1 |
| Standard private system | 0.52 (0.17, 1.57) | 1.36 (0.51, 3.67) |
| Continuity of midwifery care | 1.33 (0.64, 2.76) | 1.00 (0.60, 1.67) |
| Education and research | 2.67 (0.60, 11.96) | 1.59 (0.67, 3.78) |
| Other | 4.97 (0.64, 38.39) | 1.50 (0.63, 3.60) |

Bolded values indicate $p \leq 0.05$

### Factors associated with attitudes about pre or interconception health

There were minimal associations between participant characteristics and attitudes towards PICC on univariate analysis (Fig 6), with a few exceptions. Participants who had a registered nursing qualification had a higher likelihood of agreeing with the statement that they would encounter a health status that could be managed pre-pregnancy as part of their practice (OR 3.28; 95% CI 1.24, 8.68). However, this same group were also less likely to agree that planning to conceive is a personal decision that should only be discussed when initiated by the woman (OR 0.55; 95% CI 0.35, 0.87). Participants with more than 21 years of experience were more likely to agree with the statement that midwives are primary providers of pre and interconception care (OR 3.30; 95% CI 1.38, 7.91). However, they were also less likely to agree that planning to conceive should be discussed only when initiated by the woman (OR 0.44; 95% CI 0.23, 0.83). Further, participants in non-metropolitan areas had a lower likelihood of agreeing with the statement that PICC was within the scope of practice for midwives (OR 0.43; 95% CI 0.19, 0.98), and that they encounter health statuses that could be managed pre-pregnancy (OR 0.34; 95% CI 0.13, 0.87).

**Fig 6.**
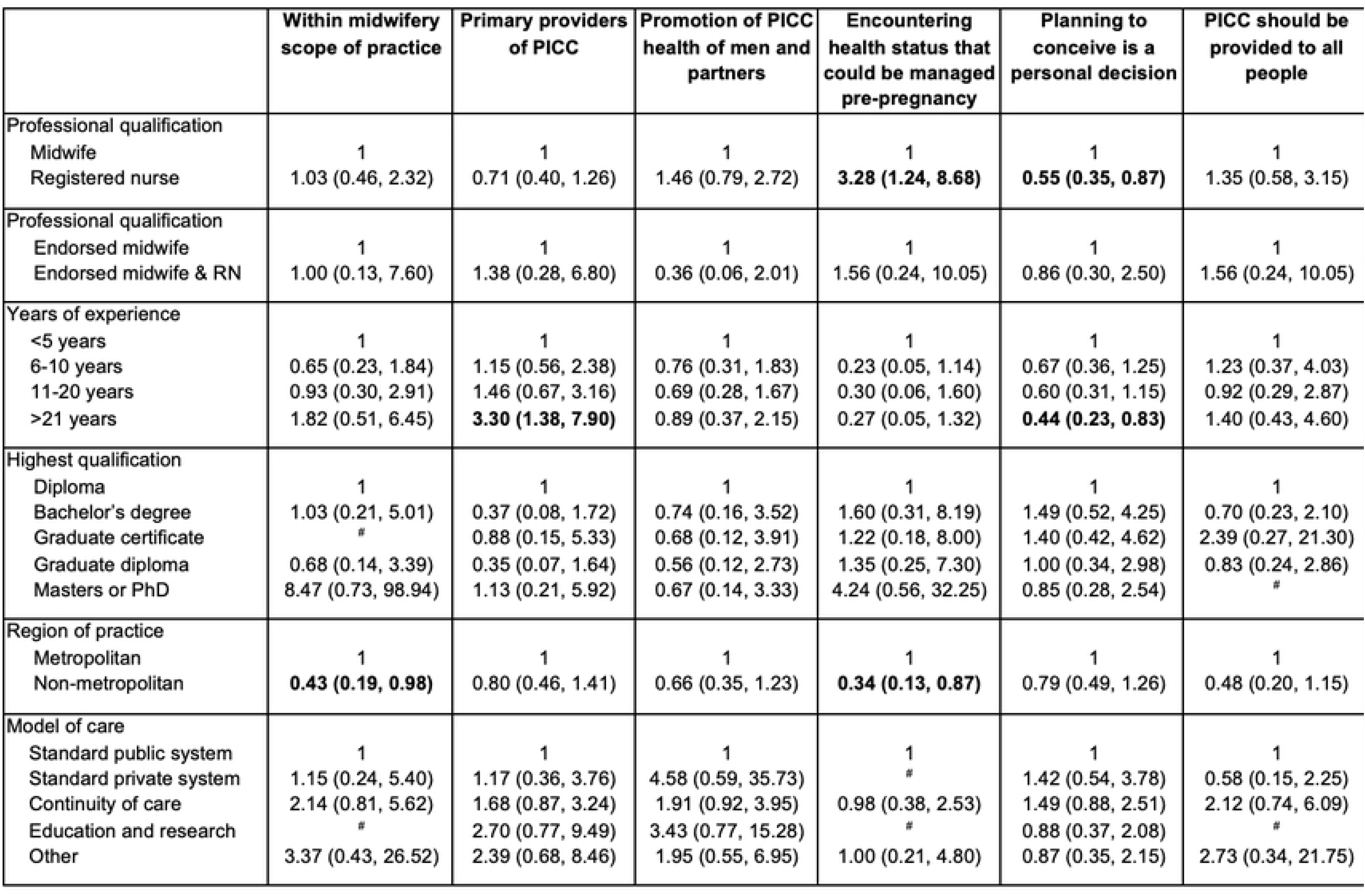
**Characteristics of participants associated with attitudes towards PICC.** Bolded values indicate *p*≤0.05 ^#^Excluded from analysis owing to low cell counts

## Qualitative results

Findings revealed three main categories i) Midwives providing PICC ii) Factors influencing midwives’ provision of PICC; and iii) Addressing learning needs. Corresponding categories were identified and are expanded on below (Fig 7).

**Fig 7.**
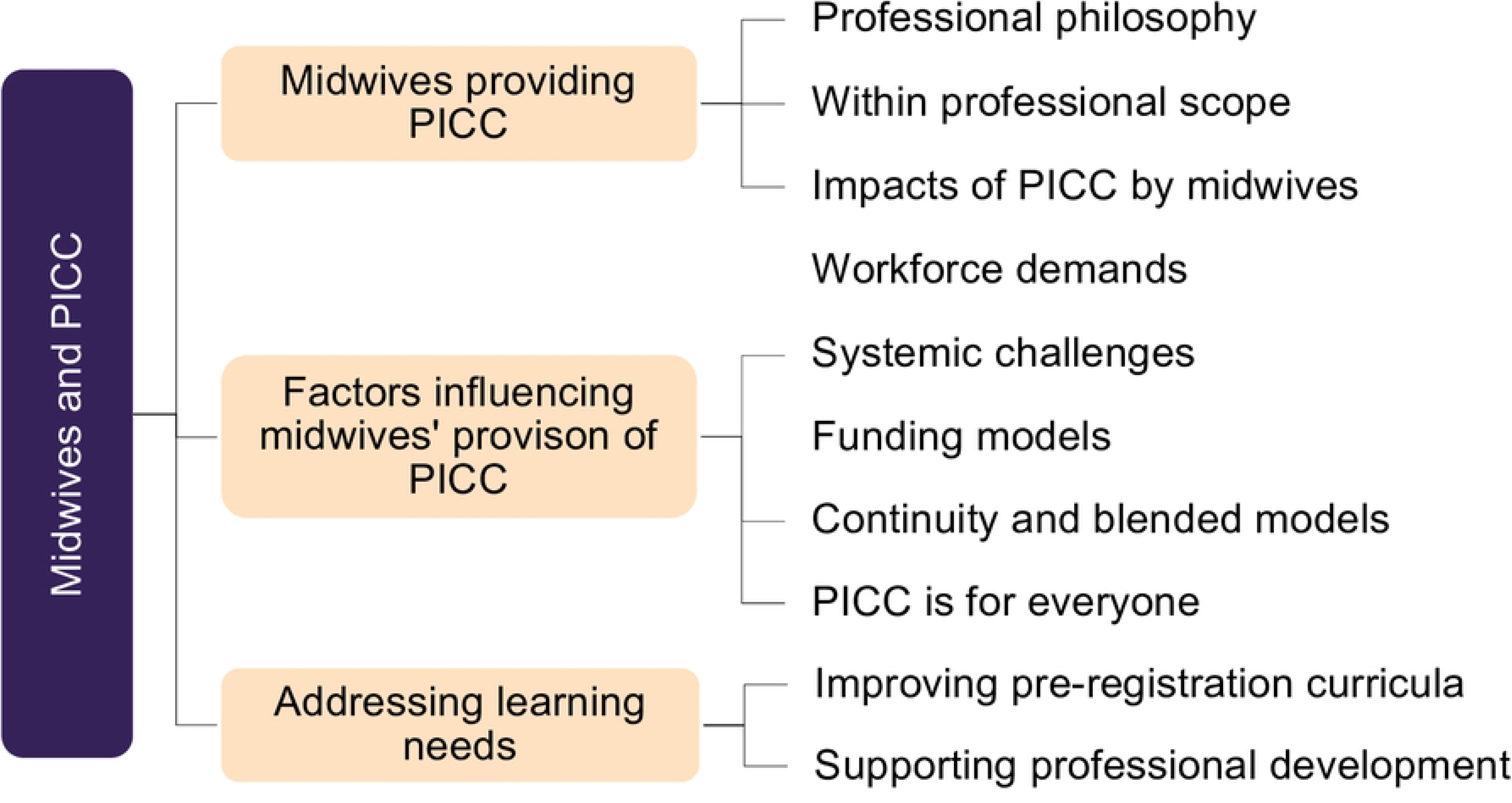
**Main categories and corresponding categories for midwives’ descriptions of preconception and interconception care.**

### 1. Midwives providing PICC

Midwives provided a range of responses indicating their existing knowledge regarding PICC. Responses included how PICC was supported by professional philosophy and scope of practice, and perceptions regarding the impact of PICC provided by midwives.

#### 1.1 Professional philosophy

There was agreement that the professional philosophy of midwifery which grounds the provision of woman-centred care, in partnership with the woman and according to her identified priorities, indicates midwives were ideally placed to provide PICC: “*Midwives are well placed to provide holistic pre/interconception care due to the profession’s underlying philosophy and emphasis on relational care”* (P232). Care provided in partnership with the woman had recognised benefits for the provision of PICC: “*Midwifery philosophy pairs perfectly with sexual health education and promotion and is so important for the provision of evidence based care that women are encouraged to be autonomous - something that is integral to midwifery practice”* (P7).

#### 1.2 Within professional scope

There was broad consensus on the provision of PICC being squarely within midwives’ professional scope: “*It makes sense to expand possibilities for midwives to provide this fundamental primary health care. It is well within our scope of practice”* (P334), with another concurring that *“pre/interconception care is very much part of the midwives’ role"* (P242). Recognition of midwives’ scope of practice across the life course confirmed the importance of midwives’ role in PICC: “*Midwives are experts… Women should have the option to access midwives from school age such as sex ed*[ucation], *actually all through the reproductive life cycle of both males and females”* (P38).

#### 1.3 Impacts of PICC by midwives

Midwives acknowledged the potential impacts of access to PICC provided by midwives on broader population health: “*Midwives should be involved in the preconception phase for all women of childbearing age to reduce the risk of morbidity and mortality for mums and babies”* (P178). The extension of positive perinatal health impacts were described as a key dividends of providing PICC education: “…*preconception care should be a priority and educating people generally to the importance of this and the positive impact it could have during the pregnancy, birth and afterwards for mother and baby”* (P246).

### 2. Factors that influence midwives’ provision of PICC

Midwives provided descriptions of barriers and enablers for the provision of PICC within their work.

#### 2.1 Workforce demands

Despite the enthusiasm around midwives’ provision of PICC some offered commentary around the pragmatics of this in the current context: “*While I think this is certainly within the scope of a midwife I can’t see the current available workforce having the numbers/time to include this in their role”* (P12). Current midwifery workforce shortages were identified as a threat to enabling midwives’ fulfilment of scope through the provision of PICC, with one participant affirming that *“… we are desperate to have a functioning midwifery workforce that can cover antenatal visits, Birth Centre, postnatal or nurseries. This needs to be addressed before we embark on any other work”* (P254).

#### 2.2 Systemic challenges

Midwives were cognisant that even though provision of PICC is within professional scope, the regulatory function of individual health services can be a barrier to midwives fulfilling their professional scope: “*The Australian public maternity care system places many limitations on the midwifery scope of practice which makes it difficult to provide this type of care”* (P232).

The current structuring of maternity services limited midwives’ ability to engage with women before or in between pregnancies: “*The truth is, seeing the women for the first time at 14-16 weeks* [gestation] *leaves a massive gap for the women to* [get] *help during the first trimester and before* [pregnancy]*. It seems it’s left up to luck if a woman has a* good [general practitioner] *that can help assist her before we take over care”* (P333). To combat this, midwives reported a commitment to providing care during ad hoc opportunities, one respondent stating: *“Where I can I am providing pre/interconception care. There is not a lot of opportunity to provide this care prior to pregnancy”* (P305).

#### 2.3 Funding models

Midwives emphasised the need for universal access with no out of pocket costs in order to achieve equitable access to PICC: “*It needs to be recognised by Medicare that it is within the scope of practice of a midwife to provide pre-conception care; therefore rebate-able* (sic) *otherwise it becomes only available to the wealthy and inaccessible to those in high need, it should be a normal part of planning a pregnancy and would greatly increase women accessing midwifery led care throughout their pregnancy and early parenting journey”* (P187). This was confirmed by a participant who relayed the challenges unique to rural areas: “[in] *rural South Australia* [women have to] *go to a GP which is costly and has severe time restraints”* (P345). Additional commentary around broader funding arrangements was provided, indicating that bundled funding allocated to the woman for her discretion to allocate would support a reorientation of care driven by individuals’ identified needs: “*Women are getting bad care because it is system based. Women should be allocated funding to access the care they choose”* (P86).

#### 2.4 Continuity and blended models improve PICC

Midwives working in continuity models reported the function of ‘extended’ postnatal care in the context of the trusting professional relationship, to ensure the woman is set up for interconception health: *“Interconception care gets provided more in continuity models in the postnatal period prior to discharge. Core staff don’t follow women up for that long, so it falls to the GP to look after”* (P270).

Similarly, midwives working in blended or hybrid professional roles indicated the utility of brief intervention education highlighting the important role that these multi-qualified professionals can fulfil: *“My best opportunities for pre/inter conception care was when working in a dual role as a midwife and child health … I was often talking to mums and dads about planning for future babies but as a midwife I only got to see people once they were already pregnant. Maternal child health space is a good place for women and midwives to connect between pregnancies”* (P6).

#### 2.5 PICC is for everyone

The role of midwives to provide PICC for all who need it was confirmed repeatedly: *“We need a big national drive to push the importance of accessing preconception health/education for BOTH woman and partner”* (P63). Another midwife described the opportunity costs to society when midwives were prevented from helping all the people in their care: *“I think it is such a shame in public care we cannot deliver care towards women’s partners. I had a situation recently where I had woman positive for chlamydia who I treated and the partner in front of me who I could not treat due to health service policy. This man was unlikely to be treated in the future due to stigma so the opportunity was likely lost, likely leading to reinfection for the woman. It is a shame we do not use these opportunities to deliver family centred primary care”* (P30).

### 3. Addressing learning needs

There was acknowledgement from participants that further education is required both at pre- and post-registration levels, with suggestions of strategies to address learning needs.

#### 3.1 Improving pre-registration curricula

There was agreement from participants that PICC should be explicitly included in pre-registration curricula, reporting that *“pre/interconception care needs to be more broadly covered in university coursework to prepare/arm newly qualified midwives with information/knowledge that has great potential to improve health outcomes for women and their babies”* (P3). Midwives acknowledged that amendments to education standards can take time but encouraged strategic approaches to embedding PICC in future version changes: *“A wide range of knowledge is required that should be updated and best evidence based. That would require planning and forward thinking”* (P27).

#### 3.2 Supporting professional development

Midwives indicated both specific areas that they would like education in, such as *“infertility management”* (P132); through to identifying a need for improved PICC knowledge more broadly: “*Preconception health care is so incredibly important … it would be amazing for midwives to have more knowledge about* [PCC] (P183). Those who had previously undertaken further/formal training indicated the utility of this in their current practice: “*I’ve valued that knowledge immensely and it has enhanced my skills and abilities as a midwife tremendously to provide family centred evidence based care in a variety of settings: community through antenatal classes, pregnancy resource centres, mothers’ groups, hospital care, antenatal clinics, global low-resource settings”* (P37).

## Discussion

This is the first study of its kind to specifically explore Australian midwives’ knowledge, attitudes and perceptions of providing PICC. Professional commentary recognising the important benefits of PICC by midwives dates back to the 1990’s (36). Global research has been undertaken, surveying a range of multidisciplinary health professionals’ perspectives and ability to provide PICC. Findings have consistently reported that midwives are ideally situated to provide PICC (37–39). Despite these international recommendations, it is noteworthy that this is the first study in Australia to explicitly ask midwives themselves about their preparedness and willingness to provide PICC.

Midwives who participated in our study provided self-scored ratings, showing most held average to excellent levels of knowledge regarding women’s health across the five domains of PICC; and those with over 11 years’ experience were three times more likely to rate in this way. This is an expected finding given the internationally established recognition of midwives being ideally placed to provide PICC due to their existing professional knowledge base, and expertise in primary reproductive care (17, 38). Given the broad strengths in PICC knowledge related to women, specific education on topics such as haemoglobin disorders, eating disorders, supplementation in pregnancy, vaccination and abortion care pathways is more likely to meet the identified professional development needs. There was agreement from midwives that pre-registration education on SRH was inconsistent and should be addressed. The midwifery education standards in Australia recognise the need for graduates to work to the international definition of the midwife, however, the provision of SRH features exclusively under the domain of postnatal care (40) which limits education about care to the interconception period only. Future versions of education standards should consider ways to further strengthen entry to registration midwifery standards for SRH including preconception care.

When considering these ratings for providing care to women’s partners/ men, findings revealed most indicated below average levels of knowledge. These results provide a clear path for areas to direct future education for midwives providing PICC. There is emerging focus on the importance of pre and interconception health for male reproductive partners as it relates to intergenerational outcomes (41, 42), and beyond these biological effects, the pre and interconception health states of all partners has lasting effects on their own health, mental health in parenthood and support provided to women within a parenting relationship (43). The organisation of PICC for men and partners requires planning, policy development and co-design (43). As conception and pregnancy is a time of opportunistic contact with healthcare for women and their partners, enhancing the knowledge and skills of midwives to provide brief interventions in this space is critical. This is reflected in both our quantitative and qualitative findings, where broadly, limited knowledge of PICC for partners and the inability to provide opportunistic healthcare to partners was raised as an area for future focus by participants.

To address knowledge gaps, midwives indicated their preference for a variety of education formats. The strong preference for online learning is unsurprising given many clinicians already receive professional education in this way (44, 45). Online webinars have become an increasingly common mode for professional development in recent years and would be supportive of the topic-based education indicated earlier. The preference for face-to-face learning from within workplaces versus from an external provider is an interesting finding and provides insight into the value and perhaps convenience and familiarity of situated, workplace learning. These findings are similar to those in a Western Australian study in which midwives emphasised their desire for face-to-face learning to facilitate not just knowledge attainment but provide practical examples on how to effectively and sensitively deliver SRH health education (23). Online webinars have become an increasingly common mode for professional development in recent years and would be supportive of the topic-based education indicated earlier. Micro-credentialed workshops and to a lesser extent, postgraduate certificate level courses were of interest to midwives and provide insight for tertiary educators or registered training organisations who might be seeking to provide these services.

Another important finding was the consistent assertion by midwives in our study that PICC must be accessible and equitably provided to all. Participants viewed PICC as not just an individual intervention, but as having valuable benefits to society as a whole. The spontaneous offering of these comments by midwives in our study exemplifies the value and potential of midwives’ voices in conversations around PICC as a critical public health strategy (46). While the positioning of midwifery care in a public health framework and the importance of midwives’ public health role has been globally asserted (17) and explored in research conducted in the United Kingdom (47, 48), further research is required to explore how midwives in Australia understand their contribution to public health. The broad interest in community-based opportunities expressed by midwives in our study also presents an opportunity for the development of innovative services where midwives can meaningfully fulfil professional scope and enact their public health role in PICC.

Harnessing and addressing the identified factors that influence midwives’ provision of PICC is essential to improving women’s access to the recognised expertise and full scope of midwifery care across the reproductive life course. Removing systemic barriers to ensure all health professionals are able to fulfil scope and provide universal access to high quality care is a recognised workforce retention strategy (49), is a key focus of national midwifery advocacy (50, 51); and is an identified priority from recent Commonwealth Government Reports (52, 53). Midwives in this study identified the need for reform in Australian healthcare funding models to ensure all women, of all backgrounds, have universal access to PICC. The need for reform was also reported in a recent national senate enquiry focused on improving women’s access to reproductive health care; recommending that midwives should have increased access to Medicare (commonwealth primary health) funding beyond the current model that limits care during pregnancy only (25). At jurisdictional levels, state governments should expand community-based, universal access to full-scope, pre and interconception care by midwives for all individuals. Such innovations would reduce health inequities and improve maternal and neonatal health outcomes [57].

The strengths of this study lie in the national inter-sectoral approach inviting participation from midwives working in all models and settings across the country which has provided important benchmarking data. Convenience sampling techniques are often employed in cross-sectional studies and have recognised limitations including the possibility of obtaining responses from individuals with distinctive views of the phenomenon under study. We cannot discount the fact that this may have occurred here however, the range of responses to each variable from midwives working in a variety of settings demonstrates heterogeneity. This survey was completed in English language only, however all midwives in Australia have a professional benchmark for reading and writing English which would not offer an obvious barrier to participation. We also had responses from those who speak languages other than English at home enabling participation from midwives from a variety of cultural and language backgrounds. The comprehensive demographic data provided enables readers to consider how findings might be transferrable to their individual settings.

## Conclusion

This study has presented novel insights regarding the knowledge and perspectives of midwives surrounding pre and interconception care (PICC). PICC is recognised as a critical element of sexual and reproductive health and within the midwifery scope of practice. Specific knowledge gaps have been identified and should inform pre and post registration midwifery curricula. Evidence on barriers to pre and interconception care provision provides direction for service reform and policy development to enhance access; and resource midwives’ provision of pre and interconception care to all women in Australia.

## Data Availability

All relevant data are within the manuscript and its Supporting Information files.

## Acknowledgements

We acknowledge all midwives who took the time to participate in this study.

## Supporting information

**S1 File. Survey.**

